# Diminished neutralization responses towards SARS-CoV-2 Omicron VoC after mRNA or vector-based COVID-19 vaccinations

**DOI:** 10.1101/2021.12.21.21267898

**Authors:** Henning Jacobsen, Monika Strengert, Henrike Maaß, Mario Alberto Ynga Durand, Barbora Kessel, Manuela Harries, Ulfert Rand, Leila Abassi, Yeonsu Kim, Tatjana Lüddecke, Pilar Hernandez, Julia Ortmann, Jana-Kristin Heise, Stefanie Castell, Daniela Gornyk, Stephan Glöckner, Vanessa Melhorn, Yvonne Kemmling, Berit Lange, Alex Dulovic, Julia Häring, Daniel Junker, Nicole Schneiderhan-Marra, Markus Hoffmann, Stefan Pöhlmann, Gérard Krause, Luka Cicin-Sain

## Abstract

SARS-CoV-2 variants accumulating immune escape mutations provide a significant risk to vaccine-induced protection. The novel variant of concern (VoC) Omicron (B.1.1.529) has the largest number of amino acid alterations in its Spike protein to date. Thus, it may efficiently escape recognition by neutralizing antibodies, allowing breakthrough infections in convalescent and vaccinated individuals. We analyzed neutralization activity of sera from individuals after vaccination with all mRNA-, vector- or heterologous immunization schemes currently available in Europe by *in vitro* neutralization assay at peak response towards SARS-CoV-2 B.1, Omicron, Beta and Delta pseudotypes and also provide longitudinal follow-up data from BNT162b2 vaccinees. All vaccines apart from Ad26.CoV2.S showed high levels of responder rates (93-100%) towards SARS-CoV-2 wild-type, but some reductions in neutralizing Beta and Delta VoC pseudotypes. The novel Omicron variant had the biggest impact, both in terms of response rates and neutralization titers. Only mRNA-1273 showed a 100% response rate to Omicron and induced the highest level of neutralizing antibody titers, followed by heterologous prime-boost approaches. Homologous BNT162b2 vaccination or vector-based AZD1222 or Ad26.CoV2.S performed less well with peak responder rates of 33%, 50% and 9%, respectively. However, Omicron responder rates in BNT162b2 recipients were maintained in our six month longitudinal follow-up indicating that individuals with cross-protection against Omicron maintain it over time. Overall, our data strongly argues for urgent booster doses in individuals who were previously vaccinated with BNT162b2, or a vector-based immunization scheme.

## 1. Introduction

Since its emergence in late 2019, SARS-CoV-2 has caused a pandemic with more than 270 million confirmed infections and more than 5 million deaths (1). While a series of vaccines have been developed with unprecedented speed and were successfully deployed to limit the burden of COVID-19, it became quickly apparent that novel SARS-CoV-2 variants had evolved, mainly in areas of high virus prevalence. Those emerging variants have accumulated mutations in the surface-exposed Spike protein, which increase virus transmissibility or promote evasion from the host immune response (2-4). Immune escape was most pronounced in SARS-CoV-2 variants Beta (B.1.351) and the currently globally dominating Delta (B.1.617.2), at least until recently. However, the November 2021 emergence of the variant B.1.1.529 (Omicron) in South Africa has raised strong concerns as its unusually high number of amino acid alterations in the Spike protein will likely contribute to an increased reinfection risk or breakthrough infections following vaccination (5). By now, a series of studies using samples from convalescent and vaccinated individuals have addressed the impact of Omicron on vaccination or infection-induced antibody neutralization, using either live-, pseudovirus neutralization or *in vitro* binding assays (6-14). These studies have shown clear losses of neutralization capacity against the Omicron variant but did not comprehensively address antibody responses in various vaccination regimens or over time. In contrast, we provide here a comprehensive assessment of vaccination schemes approved in the European Union and the UK, using an Omicron, Beta, Delta or wild-type (B.1) pseudo-neutralization assay at peak response after approximately four weeks and in a longitudinal six month follow-up for BNT162b2.

## 2. Methods

### 2.1 Sample collection and ethics statement

Serum samples analysed in this study originate from vaccinated participants of the multi-local and serial cross-sectional prevalence study on antibodies against SARS-CoV-2 in Germany (MuSPAD) study, a population-based SARS-CoV-2 seroprevalence study in eight regions of Germany from July 2020 to August 2021. The study was approved by the Ethics Committee of the Hannover Medical School (9086_BO_S_2020) and was in line with the Declaration of Helsinki. Briefly, MuSPAD is a successive cross-sectional study where certain locations were sampled longitudinally within a 3-4 month interval (15). Recruitment of eligible participants (>18 years) was based on age- and sex-stratified random sampling with information provided by the respective local residents’ registration offices. Basic sociodemographic data and information on pre-existing medical conditions including a previous SARS-CoV-2 infection or vaccination are self-reported and were documented with the Research system PIA (Prospective Monitoring and Management-App) at the study center. Peripheral blood was obtained by venipuncture using a serum gel S-Monovette (Sarstedt) and further processed according to the manufacturer`s instructions. Serum was then aliquoted at the German Red Cross Institute of Transfusion Medicine and Immunohematology and transported on dry ice to the Hannover Unified Biobank for long-term storage.

For this study, we selected samples from 82 vaccinees from the available MuSPAD sample pool to contain mRNA, vector and heterologous immunization schemes. Selection was primarily based on a consistent 21-35 day ΔT range from a complete vaccination until sampling with comparable age and gender distribution between vaccination schemes, if available paired longitudinal follow-up samples were selected of samples taken at peak response. None of the donors reported a positive SARS-CoV-2 antigen or PCR test result when questioned at the study center at the time of the blood draw. Additionally, all collected serum samples were non-reactive for nucleocapsid-specific IgG as determined with a previously published multiplex immunoassay MULTICOV-AB that contains SARS-CoV-2 Spike and nucleocapsid antigens (Supplementary Table S3, (16)), therefore excluding confounders due to infections superposed on vaccination in our cohort. Vaccination details with basic sociodemographic information and pre-existing conditions such as hypertension, cardiovascular disease, diabetes, lung disease, immunosuppression or cancer of participants are provided in Table 1 and Supplementary Table S1. As controls, the first WHO International Standard for human anti-SARS-CoV-2 immunoglobulin (code: 20/136) from the National Institute for Biological Standards and Control (NIBSC) or pre-pandemic sera samples from an anonymized Hepatitis A and Influenza virus vaccination response study at the Helmholtz Centre for Infection Research in 2014 (Hannover Medical School Ethics Committee approval number 2198-2021) were used.

**Table 1.**
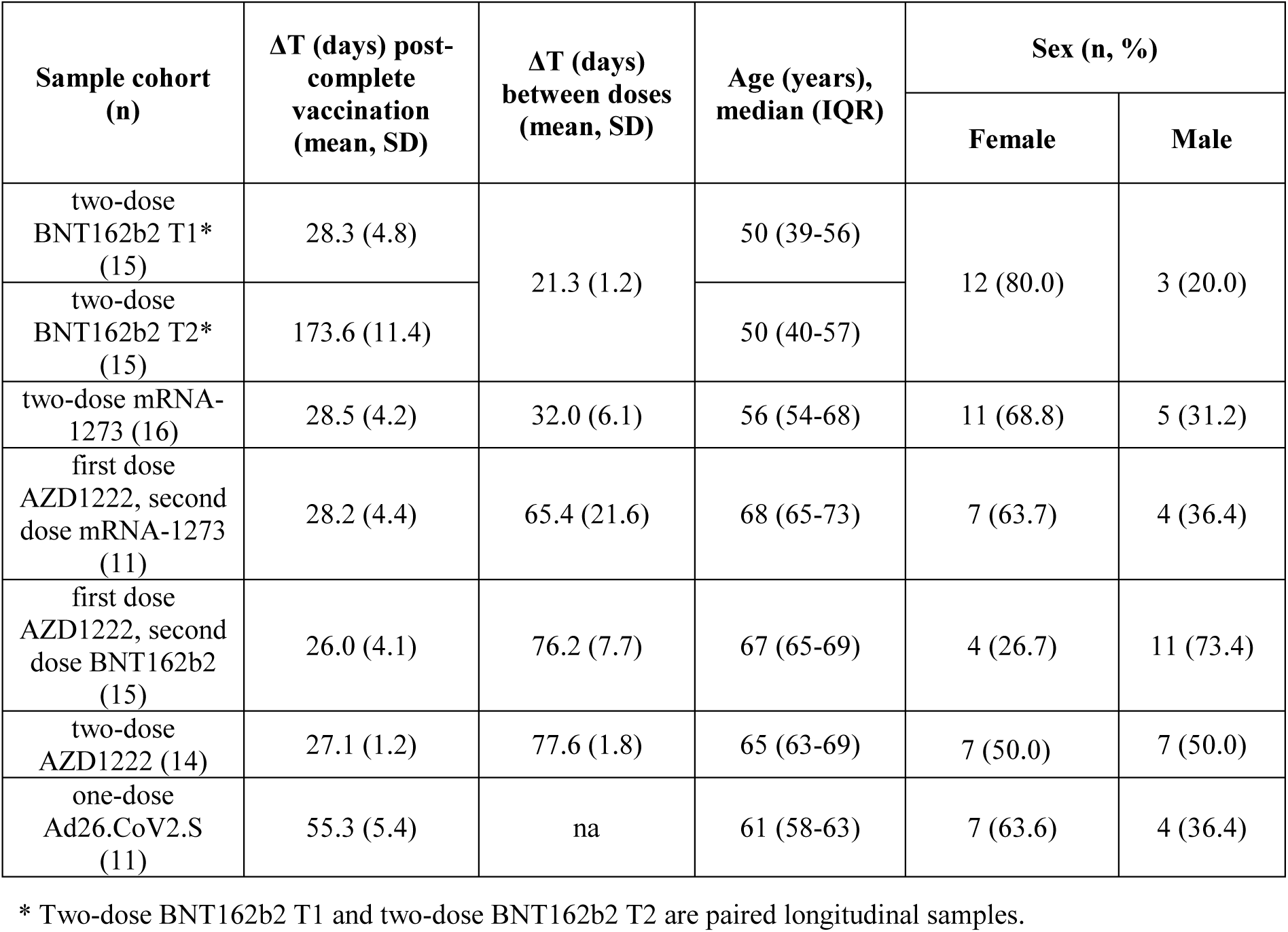
**Sample characteristics** (na: not applicable)

### 2.2 Cell culture

Vero E6 (ATCC CRL-1586), and 293T (DSMZ ACC-635) were maintained in DMEM medium supplemented with 10% fetal bovine serum (FBS), 2 mM L-glutamine, 100 U/ml penicillin and 100 μg/ml streptomycin at 37 °C in a 5% CO_2_ atmosphere. All cell lines used within this study were below a passage of 50 and were regularly checked for mycoplasma contamination. Transfection of 293T cells was performed using calcium-phosphate.

### 2.3 Plasmids

Plasmids encoding SARS-CoV-2 Spike B.1 (human codon optimized, 18 amino acid truncation at C-terminus) and SARS-CoV-2 spike of Beta (B.1.351) and Delta (B.1.617.2) have been previously reported (17-19). The expression vector for SARS-CoV-2 Spike of Omicron (based on isolate hCoV-19/Botswana/R40B58_BHP_3321001245/2021; GISAID Accession ID: EPI_ISL_6640919) was generated by Gibson assembly (13). All plasmids were sequence-confirmed by Sanger sequencing prior to use. Supplementary Table S2 provides an overview of Spike protein amino acid mutations used for SARS-CoV-2 pseudotype construction compared to the parental strain B.1.

### 2.4 Pseudotyping

Generation of rhabdoviral pseudotypes harboring SARS-CoV-2 Spike proteins was performed as described (20). In brief, 293T cells were transfected with pCG1 plasmids expressing different SARS-CoV-2 Spike proteins, using calcium-phosphate. 24 h post transfection, cells were infected with a replication-deficient reporter VSV-G (VSV*ΔG-Fluc) at an MOI of 3 for 1 h at 37 °C (21). Cells were washed once with PBS and medium containing anti-VSV-G antibody (culture supernatant from L1-hybridoma cells) was added to neutralize residual input virus. The cell culture supernatant was harvested after 16 hours, and cellular debris was removed by centrifugation at 2000 g for 5 min at 4 °C. Aliquots were stored at -80°C until use.

### 2.5 Neutralization assay

For pseudovirus neutralization, serum samples and controls were heat-inactivated at 56 °C for 30 min. Thawed samples and controls were stored at 4°C for no longer than 48 hours, prior to use. In a 96-well microtiter plate, serum samples were 2-fold serially diluted in cell culture medium (DMEM, 5 % FBS, 1% P/S, 1% L-Glu) with a dilution range of 1:10 to 1:5120. Pre-diluted samples were incubated with an equal volume of Spike protein-bearing viral particles (approximately 200 - 500 ffu/well) at 37 °C for 1 h. After incubation, the sample-virus mixture was transferred to VeroE6 cells at 100% confluence which were seeded the day before. Cells were incubated at 37 °C for 24±2 h, while infected cells were visualized using an IncuCyte S3 (Sartorius) performing whole-well scans (4x) in phase contrast and green fluorescence settings. Automated segmentation and counting of fluorescent foci defined as GFP^+^-single cells was performed using the IncuCyte GUI software (versions 2019B Rev1 and 2021B). Raw data were plotted in GraphPad prism version 9.0.2 and FRNT_50_ was calculated with a variable slope, four-parameter regression analysis. Non-responders were defined as subjects with undetectable neutralization titers at an initial serum dilution of 1:10. FRNT_50_ values of those individuals were arbitrarily set to 1. All experiments were performed with internal standard controls (pool of all tested sera), negative controls and virus-only controls to assess the nominal virus input for every single measurement.

### 2.6 Data analysis and statistics

Initial results collation and matching to metadata was done in Excel 2016 and R 4.1.0. Graphs and statistical calculations were performed using GraphPad Prism version 9.0.2 for Windows (GraphPad Software). For analysis of neutralization assay results, a Shapiro-Wilk test was used to determine normality. Focus Reduction Neutralization titer with a 50% neutralization cut-off (FRNT_50_) was calculated using a four-parameter regression analysis function. FRNT_50_ values from non-responders were set to 1 for graphical presentation only. A non-parametric Friedman’s test followed by Dunn’s multiple comparison analysis was used to compare neutralization results to different viruses in a pair-wise manner for matched samples. Two-tailed Wilcoxon matched-pairs signed rank test was used to compare neutralization of longitudinal results. A p-value of less than 0.05 was considered statistically significant.

### 2.7 Role of the funders

This work was financially supported by the Initiative and Networking Fund of the Helmholtz Association of German Research Centres through projects “Virological and immunological determinants of COVID-19 pathogenesis – lessons to get prepared for future pandemics (KA1-Co-02 “COVIPA”) to LCS and grant number SO-96 to GK, by the Deutsche Forschungsgemeinschaft (DFG, German Research Foundation) under Germany’s Excellence Strategy – EXC 2155 “RESIST” – Project ID 39087428 to LCS, and intramural funds of the Helmholtz Centre for Infection Research. SP was supported by BMBF (01KI2006D, 01KI20328A, 01KX2021), the Ministry for Science and Culture of Lower Saxony (14-76103-184, MWK HZI COVID-19) and the German Research Foundation (DFG; PO 716/11-1, PO 716/14-1). The funders had no role in study design, data collection, data analysis, interpretation, writing or submission of the manuscript. All authors had complete access to the data and hold responsibility for the decision to submit for publication.

## 3. Results

Neutralization responses towards B.1, B.1.1.529, B.1.351, and B.1.617.2 Spike-expressing rhabdoviral pseudotypes were analyzed in serum samples from 82 individuals vaccinated with either a single dose of Ad26.COV2.S, homologous two-dose BNT162b2, mRNA-1273 or AZD1222 vaccination, or heterologous AZD1222-BNT162b2 or AZD1222-mRNA-1273 vaccination at peak response, approximately four weeks after the last dose. While the WHO international standard serum showed detectable neutralization against all variants including Omicron, showing excellent sensitivity of our assay, compared to previous studies (22), pre-pandemic control sera from 2014 (n=4) showed no measurable neutralization levels (Supplementary Table S3). Neutralization potency towards Beta VoC pseudotypes were clearly reduced for all vaccination schemes, however Omicron had the strongest effect across all samples tested (Fig. 1). Vaccination with vector-based Ad26.CoV2.S performed least well (Fig. 1a), with only 73% responders against the B.1 variant, 18% for the Beta and 9% for Omicron VoC. Homologous vaccination with either AZD1222 or BNT162b2 performed better against Omicron, with 50% or 33% responders, respectively (Fig. 1b, 1c). Heterologous immunization with these two vaccines (AZD1222-BNT162b2) showed a response rate of 80% (Fig. 1d). Heterologous vaccination with AZD1222-mRNA-1273 had a similar response rate of 82% (Fig. 1e), but homologous immunization with mRNA-1273 had the highest Omicron response rate of 100% (Fig. 1f). Non-parametrical statistical comparisons showed a highly significant reduction in serum titers when Omicron neutralization was compared to B.1 for all vaccination schemes (Fig 1a-e). There was no tendency of age, sex, or pre-existing medical conditions to modify the responder status against Omicron in our cohort (Supplementary Table S3). To assess the impact of immune escape with more detail, we focused on the responders and compared geometric means of their FRNT_50_ titers (GMT). Importantly, fold-changes for groups that included non-responders are not provided in Fig. 1, because this would lead to highly artificial results and possibly over-interpretation. We therefore present the percentage of responders as primary outcome and provide GMT fold changes where calculation is reasonable (100% responders in both arms). Furthermore, for each vaccination regimen, we defined the responder subgroups (excluding non-responders defined as subjects with undetectable neutralization titers at an initial 1:10 serum dilution in either group) and compared the fold-reduction for titers that could be quantified (see Table 2). In these subsets, we observed an approximate 15-fold reduction in GMT for most vaccinations, except BNT162b2, where the reduction was 28-fold at peak response. This was consistent with the high frequency of non-responders in this subset, additionally arguing for weaker protection against the Omicron variant in this cohort.

**Fig. 1.**
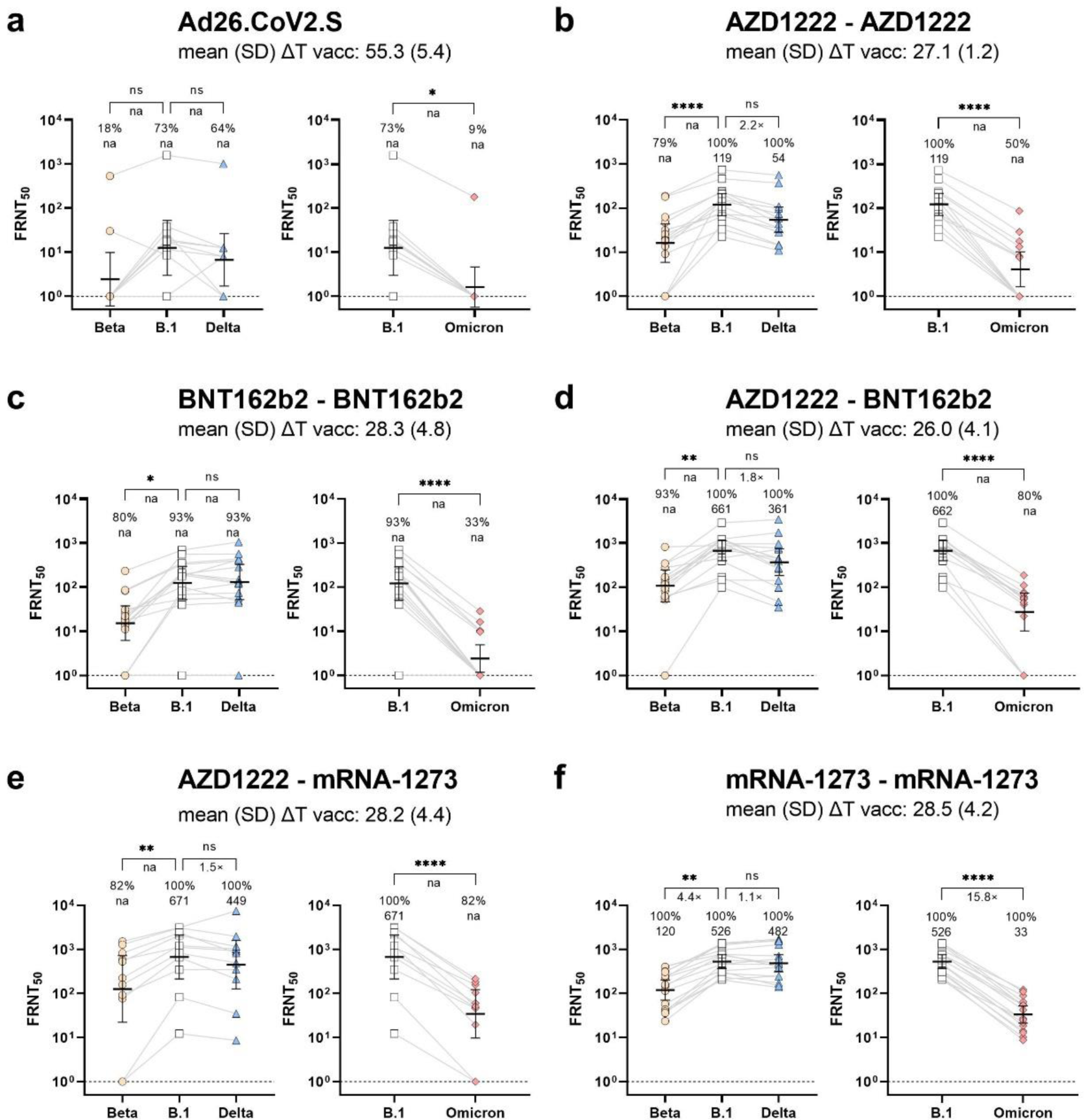
Impact of SARS-CoV-2 vaccination schemes on neutralization response towards Omicron (B.1.1.529) variant. Vaccination-induced neutralization potency against Omicron (B.1.1.529), Beta (B. 1.351), Delta (B.1.617.2) or Wuhan (B.1) pseudotypes was measured in individuals who received a vector-based vaccination with single dose Ad26.CoV2.S (n=11, a), two doses of AZD1222 (n=14, b), two doses of mRNA vaccine BNT162b2 (n=15, c), a heterologous two-dose vaccination with AZD1222-BNT162b2 (n=15, d) or AZD1222-mRNA-1273 (n=11, e), or two doses of mRNA vaccine mRNA-1273 (n=16, f) 21 to 61 days after the last dose. FRNT_50_ data is expressed for each serum sample, bold horizontal lines and whiskers are geometric means with 95% CI. Interconnecting lines represent sample data from the same donor. Non-neutralizing sample values were arbitrarily set to 1 for presentation purposes, indicated by a dashed line. Fold change in neutralization potency between SARS-CoV-2 wild-type and VoC pseudotypes is shown below p-values. Percentage (%) responder rates and FRNT_50_ geometric mean titers (GMT) per SARS-CoV-2 pseudotype are shown above the individual measurements. Fold change in neutralization potency and GMTs for SARS-CoV-2 pseudotypes are only calculated for groups where all samples had a detectable neutralizing activity, or else not applicable (na) is stated. Time between sampling and full vaccination in days is displayed as mean and SD below the vaccination scheme. Statistical analysis was performed by paired non-parametric Friedman’s test followed by a Dunn’s multiple comparison analysis. Statistical significance was defined by a value of *<0.05; ** <0.01; ***<0.001; ****<0.0001.

**Table 2.** Geometric means of responses and fold changes in Omicron responder subsets.

| Vaccination regimen | GMT (95% CI) of paired <u>responders</u> |  |  |  |
| --- | --- | --- | --- | --- |
|  | B.1 | Beta | Delta | Omicron |
| two-dose mRNA-1273 | 526.0<br>(368.1-751.5) | 119.6<br>(70.7-202.5) | 481.5<br>(70.7-202.5) | 33.2<br>(21.2-52.2) |
| first dose AZD1222,<br>second dose BNT162b2 | 902.8<br>(572.0-1425.0) | 173.9<br>(109.0-277.5) | 173.9<br>(109.0-277.5) | 61.5<br>(44.0-85.9) |
| first dose AZD1222,<br>second dose mRNA-1273 | 1323.0<br>(740.6-2363.0) | 368.9<br>(156.9-867.3) | 923.8<br>(418.5-2039.0) | 75.6<br>(41.2-138.9) |
| two-dose AZD1222 | 186.4<br>(91.2-381.3) | 46.4<br>(20.8-103.5) | 95.3<br>(41.5-232.9) | 14.6<br>(7.0-30.5) |
| one-dose Ad26.CoV2.S* | na | na | na | na |
| two-dose BNT162b2 T1<br>** | 393.0<br>(213.5-723.6) | 56.6<br>(14.3-223.5) | 503.5<br>(275.2-921.0) | 13.7<br>(7.8-24.1) |
| two-dose BNT162b2 T2<br>** | 82.6<br>(60.1-113.6) | 31.3<br>(16.8-58.2) | 85.7<br>(51.4-142.7) | 10.9<br>(7.3-16.5) |
| Vaccination regimen | Fold change in GMT of paired <u>responders</u> |  |  |  |
|  | B.1 | Beta | Delta | Omicron |
| two-dose mRNA-1273 | - | -4.4 | -1.1 | -15.8 |
| first dose AZD1222,<br>second dose BNT162b2 | - | -5.2 | -1.6 | -14.7 |
| first dose AZD1222,<br>second dose mRNA-1273 | - | -3.6 | -1.4 | -17.5 |
| two-dose AZD1222 | - | -4.0 | -1.9 | -12.8 |
| one-dose Ad26.CoV2.S* | - | na | na | na |
| two-dose BNT162b2 T1<br>** | - | -6.9 | -0.8 | -28.7 |
| two-dose BNT162b2 T2<br>** | - | -2.6 | -1.0 | -7.5 |

\* Only one subject showed detectable neutralization titers in the Omicron assay.
\*\* Two-dose BNT162b2 T1 and two-dose BNT162b2 T2 are paired longitudinal samples.

Since BNT162b2 is very commonly used, we tested the neutralization potency in BNT162b2 recipients at approximately six months post immunization as well. Similar to the peak responses, we observed a significantly weaker neutralization of the Omicron compared to the B.1 pseudotype and only 47% responders against the newly emerging VoC (Fig. 2a). Beta neutralization was slightly reduced, whereas Delta neutralization was at the same levels as B.1 at the late time point (Fig. 2a). To understand the longitudinal dynamic of humoral immunity, we used paired sera from BNT162b2 vaccine recipients at four weeks (already shown in Fig. 1c) and at six months post second dose which allowed us to compare intra-individual titer changes over time (Fig. 2b-d). While the neutralization of B.1 (Fig. 2b) and of the Delta VoC (Fig. 2d) decreased significantly over time, the time dependent reduction was less pronounced for the Beta (Fig. 2c) or the Omicron VoC (Fig. 2e). Moreover, all Omicron responders identified early after vaccination had still detectable neutralizing capacity at the late time points and two additional responders were identified in the late phase only (Fig. 2e). Therefore, the differences in neutralization titers between B.1 and Omicron responders were less pronounced at the late time points than at peak response (Table 2).

**Fig. 2.**
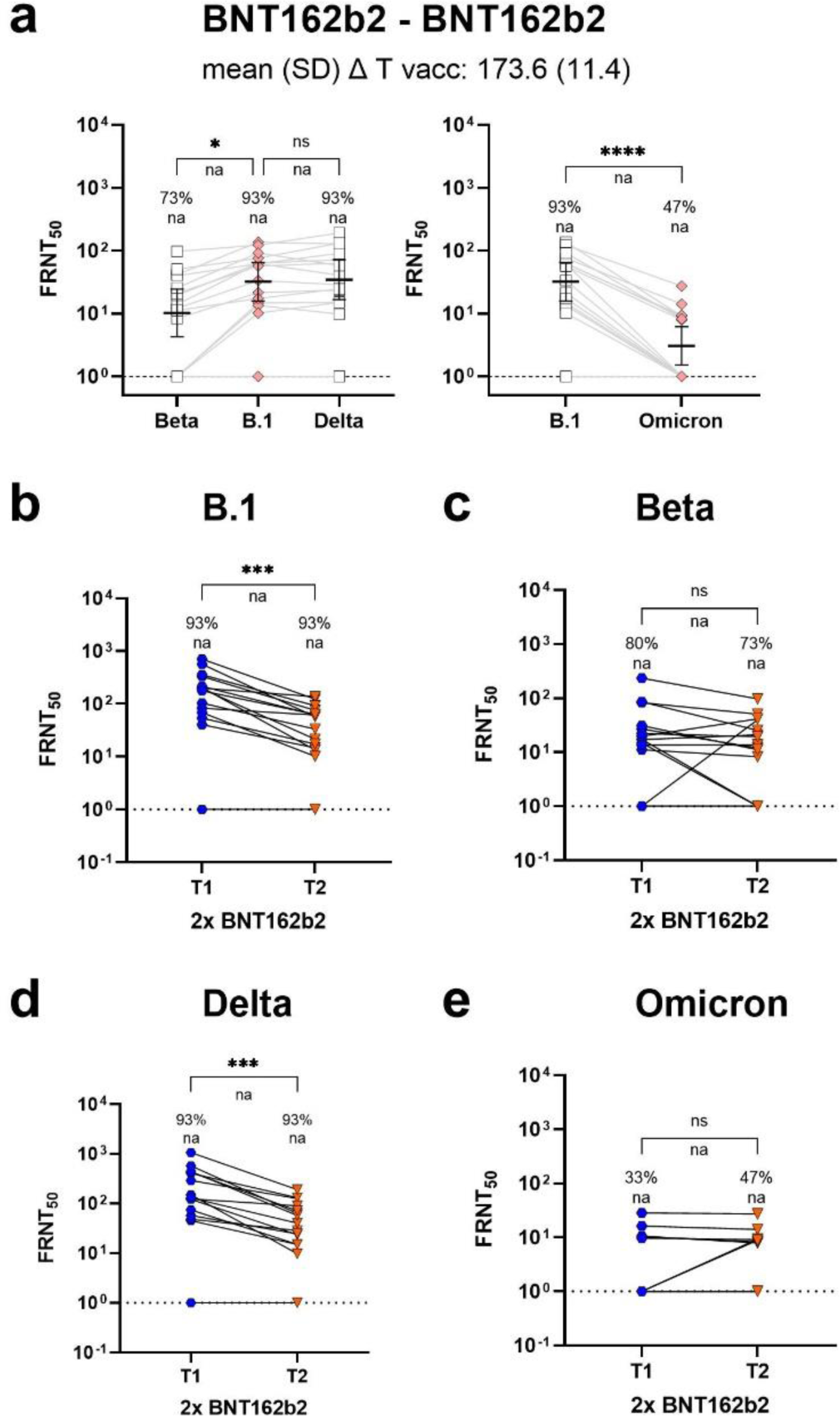
Longitudinal BNT162b2 neutralization response towards SARS-CoV-2 VoC Omicron (B.1.1.529). Neutralization capacity towards SARS-CoV-2 Omicron (B.1.1.529), Beta (B. 1.351), Delta (B.1.617.2) or Wuhan (B.1) pseudotypes was analyzed approximately six months after a two-dose BNT162b2 vaccination (n=15, a). Neutralization kinetic of paired longitudinal samples towards SARS-CoV-2 B.1 (b), Beta (c), Delta (d) and Omicron (e) pseudotypes is shown between T1 (n=15; mean (SD) ΔT after last dose: 28.3 (4.3)) and T2 (n=15; mean (SD) ΔT after last dose: 173.6 (11.4)). Peak neutralization responses of BNT162b2-vaccinated individuals from Figure 1c are displayed for clarity and comparison (b-e). FRNT_50_ data is expressed for each serum sample, bold horizontal lines and whiskers are geometric means with 95% CI. Interconnecting lines represent sample data from the same donor (a-e). Non-neutralizing samples were arbitrarily set to 1 for presentation purposes, indicated by a dashed line. Fold change in neutralization potency between SARS-CoV-2 wild-type and VoC pseudotypes is shown below p-values. Percentage (%) responder rates and FRNT_50_ geometric mean titers (GMT) per SARS-CoV-2 pseudotype are shown above the individual measurements. Fold change in neutralization potency and GMTs for SARS-CoV-2 pseudotypes are only calculated for groups where all samples had a detectable neutralizing activity, or else not applicable (na) is stated. Statistical analysis was performed by paired non-parametric Friedman’s test followed by a Dunn’s multiple comparison analysis (a) or a by two-tailed Wilcoxon matched-pairs signed rank test (b-e). Statistical significance was defined by a value of *<0.05; ** <0.01; ***<0.001; ****<0.0001.

In sum, homologous mRNA-1273 vaccination resulted in the highest responder rate, Ad26.CoV2.S in the lowest within the vaccination peak response phase, and a longitudinal follow-up showed that Omicron responses, while reduced, can be rather durable if present in the first place.

## Discussion

We provide a comprehensive overview of neutralization responses from all currently available COVID-19 vaccination schemes in the European Union and the UK not only towards the Omicron VoC, but also towards Beta and Delta VoC compared to the parental strain B.1. We expand on previous findings (11, 12) that neutralization towards Omicron is particular poor after vaccination with vector-based formulations even within the peak phase shortly after vaccination. Also consistent with other reports (13, 14), we observed a still surprisingly low cross-neutralization in BNT162b2 recipients. While some time differences exist between doses for homologous and heterologous vaccines, sampling periods after the complete scheme were within peak response making an earlier waning of BNT162b2-induced antibodies highly unlikely. While age impacts antibody titers and neutralization potency (23-25), our group of BNT162b2 recipients is slightly younger compared to the other vaccinees making age an unlikely contributor to our observation. Additionally, no tendency became apparent for gender and comorbidities either. Considering our relatively small sample size, it is however possible that this low overall response in the BNT162b2 group was a spurious observation. Nevertheless, samples showing any cross-neutralizing responses early on remained responsive to Omicron six months later. Notably, all mRNA-1273 recipients and 80% of those receiving any heterologous vaccination showed a detectable neutralization against Omicron in our analysis. It is not clear, why these vaccination protocols were more efficient against the Omicron pseudotype than BNT162b2, but it is indicative that the baseline neutralization against the B.1 pseudotype was stronger in all of them in our sample cohorts.

The detectable responsiveness to the Omicron pseudotype in all mRNA-1273 recipients differed from previous reports where usually several samples showed no measurable neutralization against Omicron (12, 14, 26). This might be due to sampling differences, a result of increased sensitivity in our assay, or both. We chose responder rates as primary outcome because this is a less biased expression than fold changes if titers from non-responsive individuals are calculated. For the same reason, we used a non-parametric assay to evaluate differences, allowing us to include samples that were below detection threshold, but obviously very low in titer. Fold changes were calculated separately on a subset of samples that showed detectable titers in all circumstances. We observed an approximately 15-fold reduction in most vaccination regimens except BNT162b2, adding evidence that Omicron cross-neutralization was impaired in this cohort. Our studies has several limitations. First, sample numbers in our cohort are low. They are however comparable to the majority of other studies to date and are well-matched on age and sex. Second, while our mean 55 day sampling time period after Ad26.CoV2.S administration is slightly longer than the four week sampling interval of the other vaccination schemes, our results are in line with others who reported poor Omicron neutralization following vaccination with Ad26.CoV2.S both at one and five months after dosing (11, 12). Third, while self-reported information about a previous SARS-CoV-2 infection or vaccination can impact study outcome, two recent publications found a 98% consistency for vaccination type and 95% for vaccination date between self-reported and administrative records or a positive predictive value of 98.2% and a negative predictive value of 97.3% between a self-reported vaccination and the detection of SARS CoV-2 antibodies (27, 28). Last, we only examine vaccination-induced antibody titers, omitting from our study people with previous infections or boosters. While these information would be valuable, we could not provide this dataset in a reasonably short time. Hence, it remains to be determined if boosters or convalescence has impacts that specifically improve neutralization efficacy by defined vaccination protocols.

Following current recommendations, a booster vaccination is generally advised after six months in many countries. Although our results do not necessarily predict failure of vaccine effectiveness and omit measuring cellular immunity or non-neutralizing antibody effects, it does however suggest that active protection against the Omicron VoC may be reduced in many vaccinated individuals. Hence, booster vaccination might be advised at earlier stages, especially for at-risk groups in the absence of a precise and clinically relevant correlate of protection.

Overall, we provide further evidence that that amino acid mutations accumulated in the B.1.1.529 Spike protein serve to escape vaccine-induced protection. In the absence of conclusive data on infectivity and disease severity, development of adapted second generation vaccinations, booster doses and careful monitoring of future variants of concern appears warranted.

## Supporting information

Supplementary Material Diminished neutralization responses towards SARS-CoV-2 Omicron VoC after mRNA or vector-based COVID-19 vaccinations

Supplementary Data Table S3 Diminished neutralization responses towards SARS-CoV-2 Omicron VoC after mRNA or vector-based COVID-19 vaccinations

## Data Availability

Raw data is provided with the manuscript. Additional data is available upon request from the corresponding authors.

## Contributors

MS, HJ, LCS conceived the study. MS, GK, BL, SC, NSM, SP and LCS procured funding. GK, BL, MS, MaH and DG designed the population-based cohort this study is based on. HJ, MS and LCS designed the experiments. HJ, HM, MYD, UR, LA, YK and TL performed the experiments. MHa, BK, MS, JH, MYD and HM performed data analysis. MS, HJ, HM, MYD generated figures and tables. MS, BK, HJ, LCS verified the underlying data. MHo, SP provided reagents. AD, DJ, JH, MS, BK, PH, BL, SC, MHa, DG, SG, JKH, VM, YvK were involved in sample or data collection and project administration. MS, HJ, LCS wrote the manuscript. All authors critically reviewed and approved the final manuscript.

## Declaration of Interest

NSM was a speaker at Luminex user meetings in the past. The Natural and Medical Sciences Institute at the University of Tübingen is involved in applied research projects as a fee for services with the Luminex Corporation. The other authors declare no competing interest.

## Acknowledgments

We want to thank again all parties involved in any capacity in making MuSPAD happen. Most of all the study participants for their willingness and commitment to make this study possible and all colleagues at the HZI, the Hannover Unified Biobank and the DRK Institute of Transfusion Medicine and Immunohematology who contributed to project administration, organization and sample processing. We thank Daniela Lenz, Ayse Barut, Inge Hollatz-Rangosch and Fawad Khan for excellent technical assistance during this study.

## Notes

### Author Declarations

The study was approved by the Ethics Committee of the Hannover Medical School (9086_BO_S_2020).

