## Supplementary Material Diminished neutralization responses towards SARS-CoV-2 Omicron VoC after mRNA or vector-based COVID-19 vaccinations for "Diminished neutralization responses towards SARS-CoV-2 Omicron VoC after mRNA or vector-based COVID-19 vaccinations"

**Supplementary Table S1.** Comorbidities of study participants (NA: not available; CVD: cardiovascular disease

| Sample cohort (n) | Comorbi-dities (n, %) | CVD (n, %) | Lung disease (n, %) | Diabetes (n, %) | Cancer (n, %) | Immuno-supp-ression (n, %) | Hyper-tension (n, %) |
|---|---|---|---|---|---|---|---|
| two-dose BNT162b2 T1* (15) | 0:9 (64.3)<br>1:4 (28.6)<br>2:1 (7.1)<br>>2: 0 (0)<br>1 NA | 0(0) | 1 (7.1) | 0 (0) | 0 (0) | 0 (0) | 5 (35.7) |
| two-dose BNT162b2 T2* (15) | 0: 8 (53.3)<br>1: 5 (33.3)<br>2: 2 (13.3)<br>>2: 0 (0) | 1 (6.7) | 1 (6.7) | 0 (0) | 0 (0) | 2 (13.3) | 5 (33.4) |
| two-dose mRNA-1273 (16) | 0: 12 (75.0)<br>1: 4 (25.0)<br>2: 0 (0)<br>>2: 0 (0) | 1 (6.3) | 1 (6.3) | 0 (0) | 1 (6.3) | 0 (0) | 1 (6.3) |
| first dose AZD1222, second dose mRNA-1273 (11) | 0: 7 (63.6)<br>1: 3 (27.3)<br>2: 1 (9.1)<br>>2: 0 (0) | 2 (18.2) | 0 (0) | 0 (0) | 0 (0) | 0 (0) | 3 (27.3) |
| first dose AZD1222, second dose BNT162b2 (15) | 0: 8 (53.3)<br>1: 3 (20.0)<br>2: 3 (20.0)<br>>2: 1 (6.7) | 1 (6.7) | 1 (6.7) | 2 (13.3) | 1 (6.7) | 0 (0) | 7 (46.7) |
| two-dose AZD1222 (14) | 0: 7 (50.0)<br>1: 6 (42.9)<br>1: 1 (7.1)<br>>2: 0 (0) | 0 (0) | 0 (0) | 2 (14.3) | 0 (0) | 0 (0) | 6 (42.9) |
| one-dose Ad26.CoV 2.S (11) | 0: 7 (63.6)<br>1: 3 (27.3)<br>2: 0 (0)<br>>2: 1 (9.1) | 1 (9.1) | 1 (9.1) | 1 (9.1) | 0 (0) | 1 (9.1) | 4 (36.4) |

* Two-dose BNT162b2 T1 and two-dose BNT162b2 T2 are paired longitudinal samples.

**Supplementary Table S2.** Amino acid mutations of Spike proteins used for SARS-CoV-2 pseudotype construction compared to the parental strain B.1. Shared mutations among the constructs are highlighted in bold.

| **B.1.351 (Beta) (EPI_ISL_700428)** | **B.1.617.2 (Delta) (EPI_ISL_1921353)** | **B.1.1.529 (Omicron) (EPI_ISL_6640919)** |
|---|---|---|
| | T19R | |
| | | A67V |
| | | 69-70del |
| D80A | | |
| | | T95I |
| | **G142D** | **G142D** |
| | | 143-145del |
| | E156G | |
| | 157-158del | |
| | | N211del/L212I |
| | | Ins214EPE |
| 242-244del | | |
| R246I | | |
| | | G339D |
| | | S371L |
| | | S373P |
| | | S375F |
| K417N | | K417N |
| | | N440K |
| | | G446S |
| | L452R | |
| | | S477N |
| | **T478K** | **T478K** |
| **E484K** | | **E484A** |
| | | Q493R |
| | | G496S |
| | | Q498R |
| **N501Y** | | **N501Y** |
| | | Y505H |
| | | T547K |
| **D614G** | **D614G** | **D614G** |
| | | H655Y |
| | | N679K |
| | **P681R** | **P681H** |
| A701V | | |
| | | N764K |
| | | D796Y |
| | | N856K |
| | D950N | |
| | | Q954H |
| | | N969K |
| | | L981F |
